# Selection of long COVID symptoms influences prevalence estimates in a prospective cohort

**DOI:** 10.1101/2022.11.09.22282120

**Authors:** Elke Wynberg, Godelieve J. de Bree, Tjalling Leenstra, Anouk Verveen, Hugo D.G. van Willigen, Menno D. de Jong, Maria Prins, Anders Boyd, the RECoVERED Study Group

## Abstract

**Background:** Studies on long COVID differ in the selection of symptoms used to define the condition. We aimed to assess to what extent symptom selection impacts prevalence estimates of long COVID.

**Methods:** In a prospective cohort of patients who experienced mild to critical coronavirus disease 2019 (COVID-19), we used longitudinal data on the presence of 20 different symptoms to evaluate changes in the prevalence of long COVID over time when altering symptom selection.

**Results:** Changing symptom selection resulted in wide variation in long COVID prevalence, even within the same study population. Long COVID prevalence at 12 months since illness onset ranged from 39.6% (95%CI=33.4-46.2) when using a limited selection of symptoms to 80.6% (95%CI=74.8-85.4) when considering any reported symptom to be relevant.

**Conclusions:** Comparing the occurrence of long COVID is already complex due to heterogeneity in study design and population. Disparate symptom selection may further hamper comparison of long COVID estimates between populations. Harmonised data collection tools could be one means to achieve greater reproducibility and comparability of results.

## Introduction

Commonly-used definitions of long COVID, or post-acute sequelae of COVID-19 (PASC), require the presence of at least one relevant symptom for a minimum period of time [1, 2]. Although most definitions stipulate that long COVID symptoms must not be linked to an alternative diagnosis, the decision as to which complaints are considered relevant is left to the discretion of researchers and clinicians. Notwithstanding existing heterogeneity in study population and design, inconsistencies in symptom selection between studies may further influence the ability to compare long COVID prevalence estimates.

These inconsistencies in symptom selection may arise from numerous methodological differences. Some studies, for instance, choose to adopt a brief questionnaire to collect data on a limited number of symptoms[3]. Other studies record the presence of over 50 distinct symptoms to encompass the full spectrum of the long COVID phenotype[4], unsurprisingly reporting higher prevalence rates of long COVID than studies using less exhaustive symptom surveys. Long COVID is also known to exhibit patterns of remitting and relapsing symptoms[4]. However, correctly attributing recurrent episodes of non-specific symptoms to long COVID remains a methodological challenge. Indeed, researchers may differ in their choice to record prevalent (i.e., at a given moment) or persistent symptoms (i.e., those that are found for two or more consecutive visits and/or from illness onset onwards) in their definition of long COVID.

No studies to date have evaluated the extent to which these methodological differences can affect prevalence estimates of long COVID. In this study, we aimed to assess how symptom selection could impact prevalence rates of long COVID within a prospective cohort.

## Methods

RECoVERED is a prospective cohort of adults who experienced mild to critical COVID-19 [5]. Methodology has been described in more detail elsewhere[5]. Briefly, study participants were enrolled within 7 days of COVID-19 diagnosis (for community-dwelling participants) or admission to hospital (for hospitalised participants). A small number of hospitalised participants were enrolled retrospectively up to 3 months after illness onset in May and June 2020, to capture patients admitted to hospital during the first wave of COVID-19 in the Netherlands. All participants had laboratory confirmation of SARS-CoV-2 infection by reverse transcriptase polymerase chain reaction (RT-PCR). Individuals residing in a nursing home and those with mental disorders deemed likely to interfere to adherence to study procedures were excluded. Study participants were interviewed by trained study staff on the presence and duration of 20 symptoms, socio-demographic details, and past medical history during the first month of follow-up. On months 2 to 12 of follow-up, participants completed monthly online surveys on the presence (in the past month) of these same symptoms; these questionnaires were repeated at month 18 and 24 of follow-up.

To demonstrate the effect of changing symptom-based definitions of long COVID, we examined the proportion of individuals from month 3 since illness onset onwards who reported at least 1 symptom among (1) any of the 20 recorded symptoms, (2) only symptoms occurring within one month of COVID-19 onset, thus assumed to be attributable to COVID-19, (3) the four most commonly-reported long COVID symptoms (i.e., fatigue, dyspnoea, loss of smell/taste, myalgia), and (4) the four long COVID symptoms that continued to be present in at least 2 consecutive surveys (i.e., persistent symptoms). Corresponding 95% confidence intervals (CIs) for each time-point were estimated according to the method for binomial proportions[6].

RECoVERED was approved by the medical ethical review board of the Amsterdam University Medical Centres (NL73759.018.20). For the current analysis, we used follow-up data collected up to 1 June 2022.

## Results

Between 11 May 2020 and 21 June 2021, 349 participants were enrolled in RECoVERED, of whom none were vaccinated for COVID-19 at enrolment. Of these participants, 292 (*n=*83.7%) completed at least 1 symptom survey. Those who did not complete any symptom survey (*n*=57/349 [16.3%]) were excluded from the current analysis; they did not differ significantly in age, sex, BMI or COVID-19 severity from those who completed at least one survey. Over half of included participants were male (*n*=170/292, 58%) and the median age was 51 years (interquartile range, IQR=36-62) (Table 1). Participants were followed for a median of 562 days (IQR=399-689). However, median follow-up time was slightly shorter among those who had had severe/critical disease (449 days, IQR=274-698) compared to those with mild (561 days, IQR=412-658) and moderate (587 days, IQR=442-701) disease (*p*=0.02).

**Table 1.**
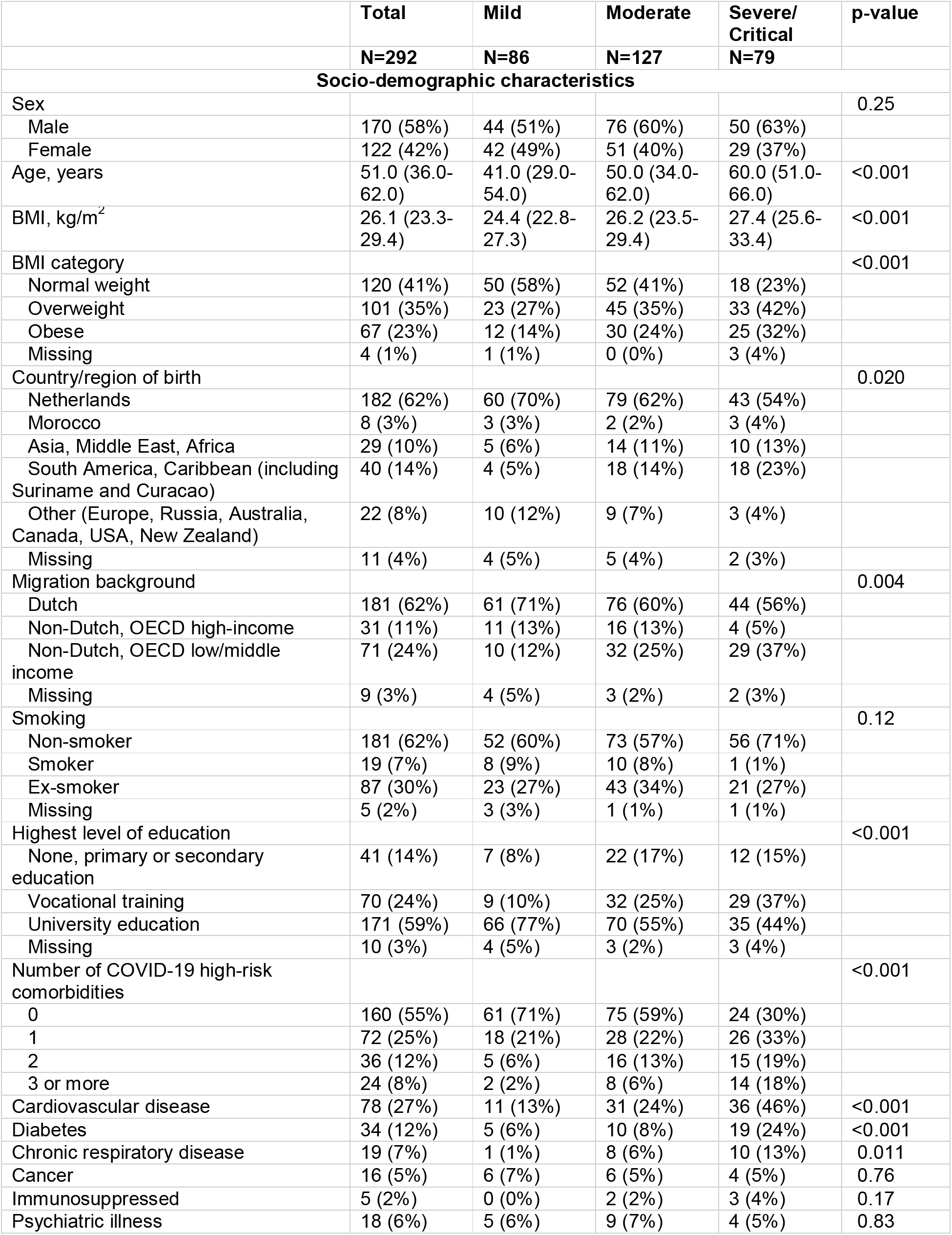
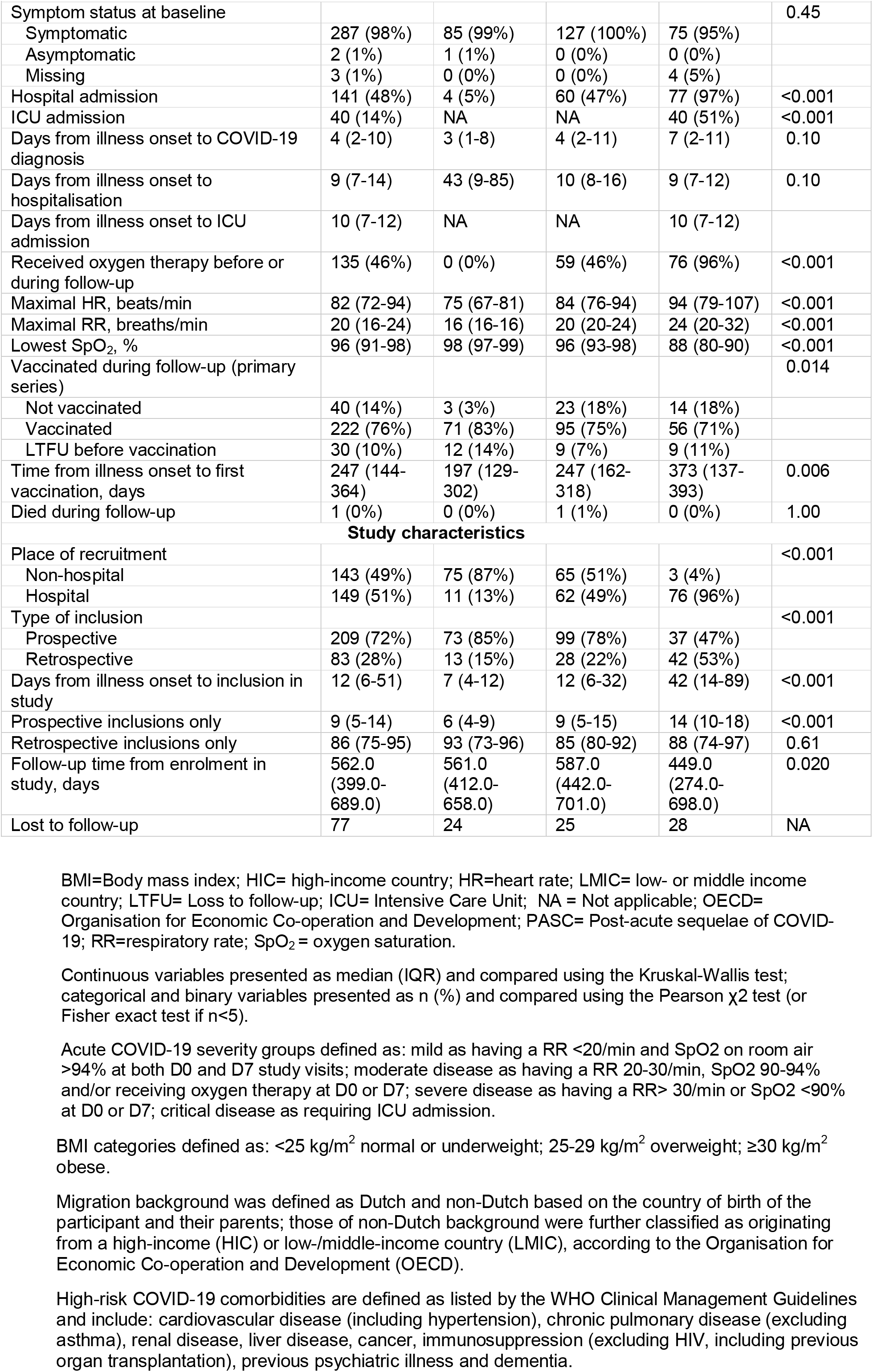
Socio-demographic, clinical and study characteristics of study participants of the RECoVERED study, Amsterdam, the Netherlands, enrolled between May 2020 and June 2021 with at least one completed symptom survey, stratified by acute COVID-19 severity

|  | Total | Mild | Moderate | Severe/<br>Critical | p-value |
| --- | --- | --- | --- | --- | --- |
|  | N=292 | N=86 | N=127 | N=79 |  |
| **Socio-demographic characteristics** |  |  |  |  |  |
| Sex |  |  |  |  | 0.25 |
| Male | 170 (58%) | 44 (51%) | 76 (60%) | 50 (63%) |  |
| Female | 122 (42%) | 42 (49%) | 51 (40%) | 29 (37%) |  |
| Age, years | 51.0 (36.0-62.0) | 41.0 (29.0-54.0) | 50.0 (34.0-62.0) | 60.0 (51.0-66.0) | <0.001 |
| BMI, kg/m<sup>2</sup> | 26.1 (23.3-29.4) | 24.4 (22.8-27.3) | 26.2 (23.5-29.4) | 27.4 (25.6-33.4) | <0.001 |
| BMI category |  |  |  |  | <0.001 |
| Normal weight | 120 (41%) | 50 (58%) | 52 (41%) | 18 (23%) |  |
| Overweight | 101 (35%) | 23 (27%) | 45 (35%) | 33 (42%) |  |
| Obese | 67 (23%) | 12 (14%) | 30 (24%) | 25 (32%) |  |
| Missing | 4 (1%) | 1 (1%) | 0 (0%) | 3 (4%) |  |
| Country/region of birth |  |  |  |  | 0.020 |
| Netherlands | 182 (62%) | 60 (70%) | 79 (62%) | 43 (54%) |  |
| Morocco | 8 (3%) | 3 (3%) | 2 (2%) | 3 (4%) |  |
| Asia, Middle East, Africa | 29 (10%) | 5 (6%) | 14 (11%) | 10 (13%) |  |
| South America, Caribbean (including Suriname and Curacao) | 40 (14%) | 4 (5%) | 18 (14%) | 18 (23%) |  |
| Other (Europe, Russia, Australia, Canada, USA, New Zealand) | 22 (8%) | 10 (12%) | 9 (7%) | 3 (4%) |  |
| Missing | 11 (4%) | 4 (5%) | 5 (4%) | 2 (3%) |  |
| Migration background |  |  |  |  | 0.004 |
| Dutch | 181 (62%) | 61 (71%) | 76 (60%) | 44 (56%) |  |
| Non-Dutch, OECD high-income | 31 (11%) | 11 (13%) | 16 (13%) | 4 (5%) |  |
| Non-Dutch, OECD low/middle income | 71 (24%) | 10 (12%) | 32 (25%) | 29 (37%) |  |
| Missing | 9 (3%) | 4 (5%) | 3 (2%) | 2 (3%) |  |
| Smoking |  |  |  |  | 0.12 |
| Non-smoker | 181 (62%) | 52 (60%) | 73 (57%) | 56 (71%) |  |
| Smoker | 19 (7%) | 8 (9%) | 10 (8%) | 1 (1%) |  |
| Ex-smoker | 87 (30%) | 23 (27%) | 43 (34%) | 21 (27%) |  |
| Missing | 5 (2%) | 3 (3%) | 1 (1%) | 1 (1%) |  |
| Highest level of education |  |  |  |  | <0.001 |
| None, primary or secondary education | 41 (14%) | 7 (8%) | 22 (17%) | 12 (15%) |  |
| Vocational training | 70 (24%) | 9 (10%) | 32 (25%) | 29 (37%) |  |
| University education | 171 (59%) | 66 (77%) | 70 (55%) | 35 (44%) |  |
| Missing | 10 (3%) | 4 (5%) | 3 (2%) | 3 (4%) |  |
| Number of COVID-19 high-risk comorbidities |  |  |  |  | <0.001 |
| 0 | 160 (55%) | 61 (71%) | 75 (59%) | 24 (30%) |  |
| 1 | 72 (25%) | 18 (21%) | 28 (22%) | 26 (33%) |  |
| 2 | 36 (12%) | 5 (6%) | 16 (13%) | 15 (19%) |  |
| 3 or more | 24 (8%) | 2 (2%) | 8 (6%) | 14 (18%) |  |
| Cardiovascular disease | 78 (27%) | 11 (13%) | 31 (24%) | 36 (46%) | <0.001 |
| Diabetes | 34 (12%) | 5 (6%) | 10 (8%) | 19 (24%) | <0.001 |
| Chronic respiratory disease | 19 (7%) | 1 (1%) | 8 (6%) | 10 (13%) | 0.011 |
| Cancer | 16 (5%) | 6 (7%) | 6 (5%) | 4 (5%) | 0.76 |
| Immunosuppressed | 5 (2%) | 0 (0%) | 2 (2%) | 3 (4%) | 0.17 |
| Psychiatric illness | 18 (6%) | 5 (6%) | 9 (7%) | 4 (5%) | 0.83 |
| Other comorbidities | 64 (22%) | 11 (13%) | 33 (26%) | 20 (25%) | 0.054 |
|  | **Total** | **Mild** | **Moderate** | **Severe/**<br>**Critical** | **p-value** |
|  | **N=292** | **N=86** | **N=127** | **N=79** |  |
| **Clinical characteristics** |  |  |  |  |  |
| Symptom status at baseline |  |  |  |  | 0.45 |
| Symptomatic | 287 (98%) | 85 (99%) | 127 (100%) | 75 (95%) |  |
| Asymptomatic | 2 (1%) | 1 (1%) | 0 (0%) | 0 (0%) |  |
| Missing | 3 (1%) | 0 (0%) | 0 (0%) | 4 (5%) |  |
| Hospital admission | 141 (48%) | 4 (5%) | 60 (47%) | 77 (97%) | <0.001 |
| ICU admission | 40 (14%) | NA | NA | 40 (51%) | <0.001 |
| Days from illness onset to COVID-19 diagnosis | 4 (2-10) | 3 (1-8) | 4 (2-11) | 7 (2-11) | 0.10 |
| Days from illness onset to hospitalisation | 9 (7-14) | 43 (9-85) | 10 (8-16) | 9 (7-12) | 0.10 |
| Days from illness onset to ICU admission | 10 (7-12) | NA | NA | 10 (7-12) |  |
| Received oxygen therapy before or during follow-up | 135 (46%) | 0 (0%) | 59 (46%) | 76 (96%) | <0.001 |
| Maximal HR, beats/min | 82 (72-94) | 75 (67-81) | 84 (76-94) | 94 (79-107) | <0.001 |
| Maximal RR, breaths/min | 20 (16-24) | 16 (16-16) | 20 (20-24) | 24 (20-32) | <0.001 |
| Lowest SpO<sub>2</sub>, % | 96 (91-98) | 98 (97-99) | 96 (93-98) | 88 (80-90) | <0.001 |
| Vaccinated during follow-up (primary series) |  |  |  |  | 0.014 |
| Not vaccinated | 40 (14%) | 3 (3%) | 23 (18%) | 14 (18%) |  |
| Vaccinated | 222 (76%) | 71 (83%) | 95 (75%) | 56 (71%) |  |
| LTFU before vaccination | 30 (10%) | 12 (14%) | 9 (7%) | 9 (11%) |  |
| Time from illness onset to first vaccination, days | 247 (144-364) | 197 (129-302) | 247 (162-318) | 373 (137-393) | 0.006 |
| Died during follow-up | 1 (0%) | 0 (0%) | 1 (1%) | 0 (0%) | 1.00 |
| **Study characteristics** |  |  |  |  |  |
| Place of recruitment |  |  |  |  | <0.001 |
| Non-hospital | 143 (49%) | 75 (87%) | 65 (51%) | 3 (4%) |  |
| Hospital | 149 (51%) | 11 (13%) | 62 (49%) | 76 (96%) |  |
| Type of inclusion |  |  |  |  | <0.001 |
| Prospective | 209 (72%) | 73 (85%) | 99 (78%) | 37 (47%) |  |
| Retrospective | 83 (28%) | 13 (15%) | 28 (22%) | 42 (53%) |  |
| Days from illness onset to inclusion in study | 12 (6-51) | 7 (4-12) | 12 (6-32) | 42 (14-89) | <0.001 |
| Prospective inclusions only | 9 (5-14) | 6 (4-9) | 9 (5-15) | 14 (10-18) | <0.001 |
| Retrospective inclusions only | 86 (75-95) | 93 (73-96) | 85 (80-92) | 88 (74-97) | 0.61 |
| Follow-up time from enrolment in study, days | 562.0 (399.0-689.0) | 561.0 (412.0-658.0) | 587.0 (442.0-701.0) | 449.0 (274.0-698.0) | 0.020 |
| Lost to follow-up | 77 | 24 | 25 | 28 | NA |

BMI=Body mass index; HIC= high-income country; HR=heart rate; LMIC= low- or middle income country; LTFU= Loss to follow-up; ICU= Intensive Care Unit; NA = Not applicable; OECD= Organisation for Economic Co-operation and Development; PASC= Post-acute sequelae of COVID-19; RR=respiratory rate; SpO<sub>2</sub>= oxygen saturation.
Continuous variables presented as median (IQR) and compared using the Kruskal-Wallis test; categorical and binary variables presented as n (%) and compared using the Pearson $\chi^2$ test (or Fisher exact test if $n < 5$ ).
Acute COVID-19 severity groups defined as: mild as having a RR <20/min and SpO<sub>2</sub> on room air >94% at both D0 and D7 study visits; moderate disease as having a RR 20-30/min, SpO<sub>2</sub> 90-94% and/or receiving oxygen therapy at D0 or D7; severe disease as having a RR > 30/min or SpO<sub>2</sub> <90% at D0 or D7; critical disease as requiring ICU admission.

Across all time-points, prevalence estimates were higher when adopting a more inclusive selection of long COVID symptoms (Figure 1). The proportion of participants with long COVID at month 3 since illness onset varied widely, from 56.0% (95%CI=48.9-62.9) when restricting to persistent key long COVID symptoms to 81.2% (95%CI=74.9-86.2) when counting any of the 20 recorded symptoms. The difference between proportions was even greater at month 12, ranging from 39.6% (95%CI=33.4-46.2) to 80.6% (95%CI=74.8-85.4).

**Figure 1.**
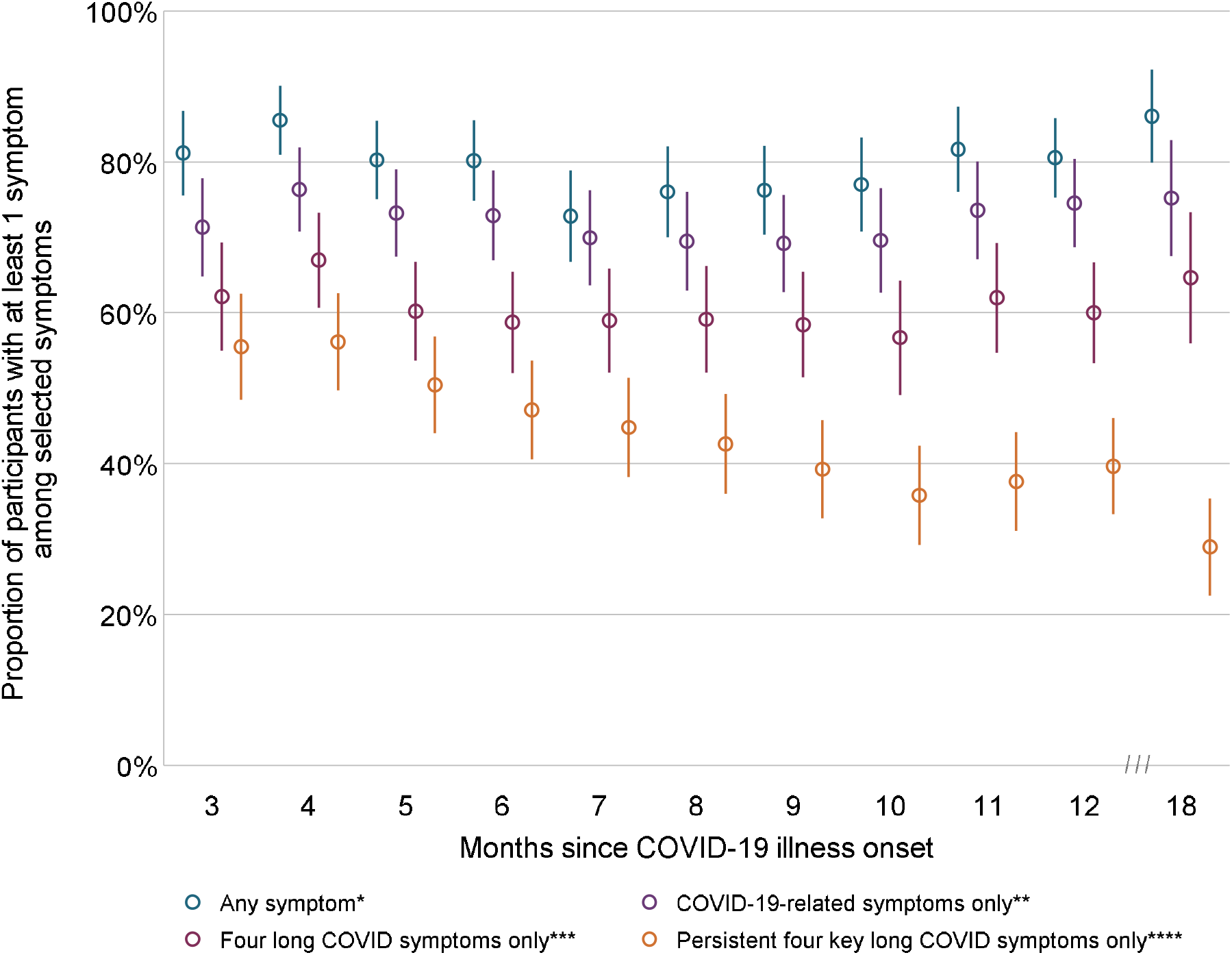
Proportions over time of RECoVERED cohort participants (Amsterdam, the Netherlands) enrolled between 11 May 2020 and 21 June 2021 with long COVID, according to four different definitions with changing selection of symptoms. Data presented above are from month 3 onwards, in line with the common definitions of long COVID. Subsequent selection of these symptoms was made according to the following criteria: * Any of the 20 symptoms included in the questionnaire ** COVID-19 symptoms, including only those symptoms with a reported start date within the first month of overall COVID-19 onset *** Counting only the four most commonly-reported long COVID symptoms in our cohort: fatigue, dyspnoea, loss of smell/taste and myalgia **** Counting only the four most commonly-reported long COVID symptoms in our cohort if they had persisted since illness onset (i.e., individuals with a later relapse in any of these four symptoms were no longer included in the numerator)

When stratifying by acute COVID-19 severity, prevalence estimates of long COVID differed most when defining long COVID according to persistent key long COVID symptoms (Supplementary Figure S1). At 18 months after illness onset, the prevalence of long COVID was 39.0% (95%CI=25.4-54.6) among those with severe/critical disease, 29.8 (95%CI=21.0-40.4) among those with moderate disease, and 21.5% (13.1-33.1) among participants with mild COVID-19, defined according to persistent key long COVID symptoms.

## Discussion

Long COVID studies to date have differed in the number and type of symptoms considered relevant. Selection of symptoms based on presumed underlying pathophysiology or persistence over time remains a subjective process, making generalisability across studies extremely challenging. We found that, even within the same cohort, the estimated prevalence of long COVID at 12 months could differ by almost 60% according to the level of stringency used to select symptoms. As expected, more restrictive methods resulted in a lower proportion of participants being identified as having at least one symptom. We also observed a lower prevalence when only participants with longitudinally persistent symptoms were considered to have long COVID, thus excluding more fleeting symptom episodes. This is likely to represent partly between-month relapsing-remitting pattern of symptoms in some long COVID patients, and demonstrate that cross-sectional estimates may be biased towards higher prevalence due to including the transient presence of non-specific symptoms.

Our observations have important downstream consequences for prevention and intervention. Firstly, particular symptoms may be correlated with specific risk factors. As a consequence, symptom selection could affect a study’s ability to identify these determinants. Moreover, variation in symptom selection may have consequences for outcome measurement in clinical trials for novel treatments for long COVID. If recovery is defined by the resolution of a restricted list of symptoms, and this list varies between clinical trials, the probability of achieving the outcome of interest may vary widely between studies. Indeed, disparities in symptom selection when defining long COVID directly impacts the definition of what we mean by recovery, adding further methodological complexity. Finally, wide variation in estimating the prevalence of long COVID may introduce confusion among the general public about the condition, and reinforce mistrust by parties who are sceptical about the physiological nature of the condition[7]. These implications may, in turn, amplify stigma towards individuals with long COVID, hinder access to care, and diminish investment in further research.

Our study’s strengths are the long follow-up time, prospective design, representation of the full spectrum of COVID-19 disease severity, and comprehensive longitudinal symptom data. This analysis also presents a novel perspective on the underlying methodology of a rapidly-growing evidence base on long COVID. However, our study also has limitations. Differential loss-to-follow-up according to COVID-19 severity may have exerted a downward bias on later prevalence estimates, with participants with severe/critical disease having a relatively shorter follow-up time than those with mild or moderate COVID-19. Interestingly, however, prevalence estimates appeared to increase slightly from month 11 after illness onset onwards in all cross-sectional estimates. This can most likely be explained by residual confounding of calendar time: those who were infected early on in the pandemic may have had reduced access to therapeutic interventions compared to those infected more recently. When stratifying by acute COVID-19 severity, this effect seemed most pronounced in those who had had mild or severe/critical disease, suggesting that clinical management for these groups has changed more over time than among those with moderate disease (Supplementary Figure S1). Finally, our symptom data was not complemented with qualitative data; a mixed-methods approach may have shed further light on which symptoms were directly associated with long COVID.

In conclusion, we have demonstrated the drastic variation in long COVID prevalence with symptom-based definitions. Standardisation of the selection of long COVID symptoms is therefore urgently needed. Harmonised data collection tools, as previously suggested [8], could be one means to achieve greater comparability of results. Nevertheless, the vast range of symptoms linked to the long COVID phenotype makes it difficult to balance comprehensiveness with reproducibility – posing the risk of over-simplification of a complex condition. Consensus from both patient and clinical societies, in combination with robust qualitative data, is crucial to ensure that long COVID is defined in a standardised but inclusive way.

## Data Availability

All data produced in the present study are available upon reasonable request to the authors.

## Statements & Declarations

## Acknowledgements

This research letter was prepared on behalf of the RECoVERED Study Group:

At the Public Health Service of Amsterdam: Ivette Agard, Jane Ayal, Floor Cavdar, Marianne Craanen, Udi Davinovich, Annemarieke Deuring, Annelies van Dijk, Ertan Ersan, Laura del Grande, Joost Hartman, Nelleke Koedoot, Romy Lebbink, Dominique Loomans, Agata Makowska, Tom du Maine, Ilja de Man, Amy Matser, Lizenka van der Meij, Maria Oud, Marleen van Polanen, Clark Reid, Leeann Storey, Marc van Wijk.

At the Amsterdam University Medical Centres: Joost van den Aardweg, Joyce van Assem, Marijne van Beek, Thyra Blankert, Maartje Dijkstra, Orlane Figaroa, Leah Frenkel, Marit van Gils, Jelle van Haga, Xiaochuan (Alvin) Han, Agnes Harskamp-Holwerda, Mette Hazenberg, Soemeja Hidad, Anja Lok, Nina de Jong, Hans Knoop, Neeltje Kootstra, Lara Kuijt, Eric Moll-van Charante, Pythia Nieuwkerk, Colin Russell, Karlijn van der Straten, Annelou van der Veen, Bas Verkaik, Gerben-Rienk Visser.

## Author contributions

Conceptualisation: Maria Prins, Anders Boyd, Tjalling Leenstra, Godelieve de Bree, Elke Wynberg. Data collection: Anouk Verveen, Hugo van Willigen, Elke Wynberg. Formal analysis and investigation: Anders Boyd, Elke Wynberg. Writing – original draft: Maria Prins, Anders Boyd, Elke Wynberg. Writing – review and editing: All authors. Funding acquisition: Maria Prins, Godelieve de Bree, Menno de Jong. Supervision and project management: Maria Prins, Godelieve de Bree, Menno de Jong, Tjalling Leenstra.

## Ethical Approval and Patient Consent Statement

RECoVERED is an observation study that was approved by the medical ethical review board of the Amsterdam University Medical Centres (NL73759.018.20). All participants of the RECoVERED study provided written informed consent.

## References

1. WHO. A clinical case definition of post COVID-19 condition by a Delphi consensus. World Health Organization (WHO) clinical case definition working group on post COVID-19 condition 2021.

2. (NICE) NIfHCE. COVID-19 rapid guideline: managing the long-term effects of COVID-19 NG188 Vol. 2021: NICE, 2021.

3. Blomberg B, Mohn KG-I, Brokstad KA, et al. Long COVID in a prospective cohort of home-isolated patients. Nature Medicine 2021; 27(9): 1607–13.

4. Tran V-T, Porcher R, Pane I, Ravaud P. Course of post COVID-19 disease symptoms over time in the ComPaRe long COVID prospective e-cohort. Nature Communications 2022; 13(1): 1812.

5. Wynberg E, van Willigen HDG, Dijkstra M, et al. Evolution of Coronavirus Disease 2019 (COVID-19) Symptoms During the First 12 Months After Illness Onset. Clinical Infectious Diseases 2021.

6. Dean N, Pagano M. Evaluating Confidence Interval Methods for Binomial Proportions in Clustered Surveys. Journal of Survey Statistics and Methodology 2015; 3(4): 484–503.

7. Rubin R. As Their Numbers Grow, COVID-19 “Long Haulers” Stump Experts. JAMA 2020; 324(14): 1381–3.

8. Sigfrid L, Cevik M, Jesudason E, et al. What is the recovery rate and risk of long-term consequences following a diagnosis of COVID-19? A harmonised, global longitudinal observational study protocol. BMJ Open 2021; 11(3): e043887.

