## Supplementary Figure S1 for "Selection of long COVID symptoms influences prevalence estimates in a prospective cohort"

### Supplementary Figure S1. Proportions of RECoVERED cohort participants (Amsterdam, the Netherlands) enrolled between 11 May 2020 and 21 June 2021 with long COVID over time, according to four definitions with changing selection of symptoms, stratified by acute COVID-19 severity

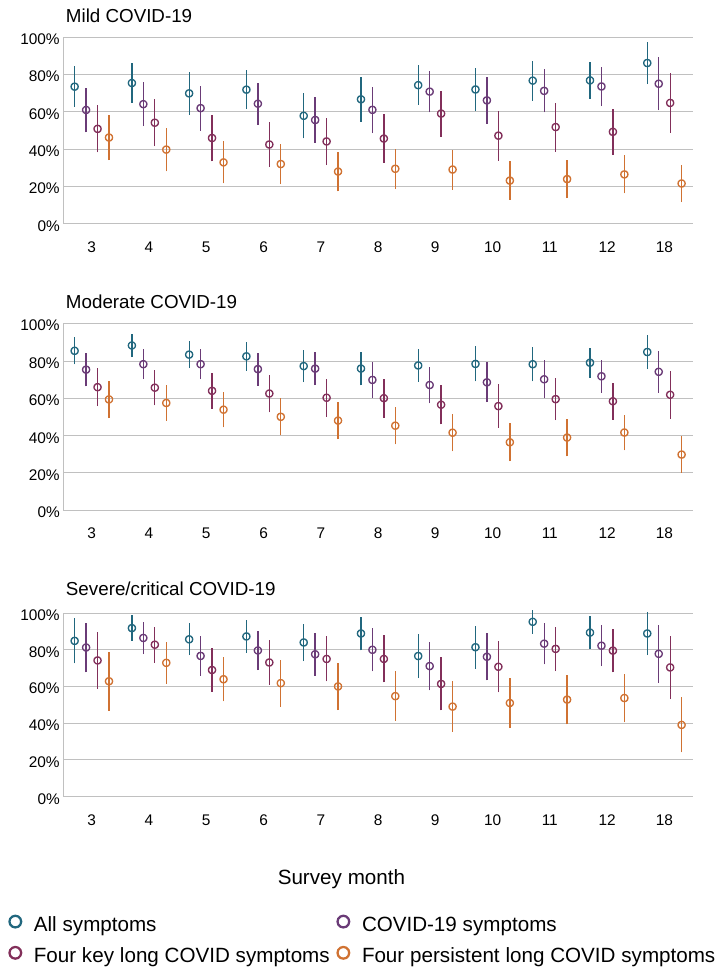

Acute COVID-19 severity groups defined as: mild as having a RR <20/min and SpO2 on room air >94% at both D0 and D7 study visits; moderate disease as having a RR 20-30/min, SpO2 90-94% and/or receiving oxygen therapy at D0 or D7; severe disease as having a RR> 30/min or SpO2 <90% at D0 or D7; critical disease as requiring ICU admission.
